# BLOod Test Trend for cancEr Detection (BLOTTED): protocol for an observational and prediction model development study using English primary care electronic health records data

**DOI:** 10.1101/2022.09.21.22280203

**Authors:** Pradeep S. Virdee, Clare Bankhead, Constantinos Koshiaris, Cynthia Wright Drakesmith, Jason Oke, Diana Withrow, Subhashisa Swain, Kiana Collins, Lara Chammas, Andres Tamm, Tingting Zhu, Eva Morris, Tim Holt, Jacqueline Birks, Rafael Perera, FD Richard Hobbs, Brian D. Nicholson

**Author notes:** **Corresponding author** Author: Brian D. Nicholson, Address: Radcliffe Primary Care Building, Radcliffe Observatory Quarter, Woodstock Road, University of Oxford, Oxford, OX2 6GG, UK. Pradeep S. Virdee (PSV), Clare Bankhead (CB), Constantinos Koshiaris (CK), Cynthia Wright Drakesmith (CWD), Jason Oke (JO), Diana Withrow (DW), Subhashisa Swain (SS), Kiana Collins (KC), Lara Chammas (LC), Andres Tamm (AT), Tingting Zhu (TZ), Eva Morris (EM), Tim Holt (TH), Jacqueline Birks (JB), Rafael Perera (RP), Richard Hobbs (RH), Brian D. Nicholson (BDN).

## Abstract

**Background:** Simple blood tests can play an important role in identifying patients for cancer investigation. The current evidence base is limited almost entirely to tests used in isolation. However, recent evidence suggests combining multiple types of blood tests and investigating trends in blood test results over time could be more useful to select patients for further cancer investigation. Such trends could increase cancer yield and reduce unnecessary referrals. We aim to explore whether trends in blood test results are more useful than symptoms or single blood test results in selecting primary care patients for cancer investigation. We aim to develop clinical prediction models that incorporate trends in blood tests to identify risk of cancer.

**Methods:** Primary care electronic health records data from the English Clinical Practice Research Datalink Aurum primary care database will be accessed and linked to cancer registrations and secondary care datasets. Using a cohort study design, we will describe patterns in blood testing (Aim 1) and explore associations between covariates and trends in blood tests with cancer using mixed-effects, Cox, and joint models (Aim 2). To build the predictive models for risk of cancer, we will use multivariate joint modelling and machine-learning, incorporating simultaneous trends in multiple blood tests, together with other covariates (Aim 3). Model performance will be assessed using various performance measures, including c-statistic and calibration plots.

**Discussion:** These models will form decision rules to help general practitioners find patients who need referral for further investigation of cancer. This could increase cancer yield, reduce unnecessary referrals, and give more patients the opportunity for treatment and improved outcomes.

## Introduction

A recent clinical review concluded that simple blood tests have an important role in identifying patients for cancer investigation^1^. However, analysis of National Cancer Diagnosis Audit in Primary Care data suggests that primary care investigations may delay referral^2^. Smarter use of blood tests to select patients for further cancer investigation could increase cancer yield and reduce unnecessary referrals, minimising the psychological and physical harm to patients and economic costs of unnecessary testing.

The current evidence base for using blood tests to identify patients at risk of cancer in primary care is almost entirely limited to single blood tests^1^. Except for anaemia and jaundice, the risk of individual cancers associated with blood test abnormalities is too low to warrant urgent cancer investigation^3^. For all cancers combined, the following blood test abnormalities increase the risk of cancer above the three percent threshold recommended by the National Institute for Health and Care Excellence (NICE) for urgent investigation: low albumin, raised platelets, raised calcium, and raised inflammatory markers^1^. Whilst these test abnormalities give general practitionors (GPs) an indication that cancer may be present, they leave uncertainty over which cancer(s) should be investigated. In practice, GPs may interpret test results in combination with other contemporaneous tests or previous results of the same test, particularly if the current result is abnormal.

Methodological innovation is required to understand whether incorporating blood test change over time may provide more accurate cancer prediction for cancer overall and for individual cancer sites. For example, a patient with a low-normal haemoglobin may not be regarded as high risk if the haemoglobin result is interpreted in isolation and an abnormal/normal binary threshold is used. However, a low-normal result following years of high-normal results may represent an opportunity for cancer investigation. Methods for incorporating repeated measures data into clinical decision rules are well known^4^. Our group has recently completed analyses investigating trends in multiple components of the full blood count blood test in the ten years prior to colorectal cancer diagnosis^5^. We also developed prediction models (using joint modelling) designed for early detection of colorectal cancer, using only data earlier than two years before diagnosis, which incorporated trends to identify two-year risk of diagnosis^6^. The sex-stratified trends indicated that a simultaneous patient-level decline in haemoglobin and mean corpuscular volume and rise in platelet count from a steady trend increased the risk of colorectal cancer diagnosis in two years from the current blood test, with good discrimination and calibration^6^. Serial testing may hold greater potential to rule-in and rule-out further cancer investigation.

In addition to serial testing, test combinations may hold greater potential to rule-in and rule-out patients for further cancer investigation^7^. A recent case-control study illustrated that a normal erythrocyte sedimentation rate plus a normal haemoglobin reduced the risk of myeloma sufficiently to rule-out the need for further investigation^8^. A cohort study of patients referred to a Danish Multidisciplinary Diagnostic Centre (MDC) showed combinations of abnormal tests markedly increased the probability of cancer being diagnosed^9^. However, multivariable cancer-risk prediction models widely accessible to National Health Service (NHS) GPs incorporate multiple symptoms and risk factors but not blood test results^10, 11^. Simple clinical scores including age-group, sex, and seven simple primary care blood tests (albumin, alkaline phosphatase, C-reactive protein, haemoglobin, liver enzymes, platelets, and total white cell count) could be used to select patients with unexpected weight loss who do and do not warrant further cancer investigation^12^. Internal validation of these risk scores have shown good discrimination between patients with and without cancer, were well calibrated at the levels of risk that decisions to investigate are made in primary care, and have shown superior clinical utility compared to models including only age, sex, and symptoms^12^. It remains unclear whether these findings are valid in external NHS and primary care datasets from abroad and whether similar scores could be developed for patients with other cancer symptoms.

There is therefore a pressing need for studies to establish the optimal and most rational use of trends in simple and commonly available blood tests to select primary care patients for urgent cancer investigation.

## Aims and objectives

The overall aim of BLOTTED is to understand whether incorporating changes in blood tests over time could optimise the selection of primary care patients for cancer investigation compared to models including symptoms, signs, and single blood test results.

### Aim 1.1

To describe results and trends of blood tests in individuals attending NHS primary care between 2000 and 2019 overall, by age, sex, ethnicity, deprivation, and comorbidity. This aim will evaluate the underlying epidemiology of blood test change over time in a primary care population.

### Aim 1.2

To describe whether changes in blood test trends occur prior to cancer symptoms, single measurement blood test abnormalities, screening findings, and referrals for suspected cancer overall and by type of blood test, symptom, referral pathway, and cancer characteristics (diagnosis route, site, grade, histology, stage). This aim will assess opportunities for earlier cancer diagnosis, such as referral for urgent cancer investigation based on changes in the trend of blood tests instead of symptoms or abnormal single blood test measurements.

### Aim 2.1

To test the association between blood test trends and subsequent cancer overall and by age, sex, and cancer characteristics (diagnosis route, site, grade, histology, stage). This aim will establish the association between cancer diagnosis and trends in blood tests in the years leading up to cancer diagnosis and identify when blood test trends could be used to prompt cancer investigation.

### Aim 2.2

To explore whether blood test trends are independent of trends in other blood tests and clinical features of cancer (symptoms, signs, and individual blood test results). This aim will examine whether blood test trends should be considered in isolation or if their predictive value increases when combined with other blood test trends and/or clinical features.

### Aim 3

To develop and validate decision rules (prediction models) to select individuals attending NHS primary care for cancer investigation. This aim will pull together the learnings from the prior aims into prediction models to derive and test a clinical strategy for GPs to guide patient selection for cancer investigation, comparing the performance of these models with performance of existing models.

## Methods

Aims 1 and 2 will be reported following the STROBE guidelines for observational research. Aim 3 will be reported following the TRIPOD guidelines for development and/or validation of prediction models.

### Data

Primary care electronic health records data will be obtained from the English Clinical Practice Research Datalink (CPRD) Aurum^13^. Cancer diagnoses will be obtained from the National Cancer Registration and Analysis Service (NCRAS), Hospital Episode Statistics (HES) database, and Office of National Statistics (ONS) (if related to death). Our blood test data preparation and reporting will follow the steps outlined in our previous work^14^.

### Study design

A cohort study will be used for each aim, allowing for an appropriate assessment of absolute risk of cancer diagnosis.

### Participants

We will include patients registered in CPRD between January 1^st^ 2000 and December 31^st^ 2019. Patients will be eligible for linkage with NDRS, HES, and ONS databases. We will exclude patients with less than 12 months registered with the general practice or less than 2 years of follow-up data following study entry.

### Outcome

An incident diagnosis of any cancer made during the study period recorded. Diagnoses will be obtained primarily from the NDRS database, with additional diagnoses obtained from the CPRD, HES, and ONS databases. Patients without a diagnosis will be censored at the earliest of date of leaving the practice, death, or 31^st^ December 2019 (data cut).

A validated library of Read/SNOMED CT/ICD-10 codes developed previously by this group will be used to identify all incident cancer diagnoses throughout the study period. Data will also be extracted on cancer site, stage, grade, and histology at diagnosis.

### Predictors

We will explore trends in the blood test results including the full blood count, liver function, and renal function tests (a full list is in Box 1):

#### Box 1

**blood tests under investigation in BLOTTED**

##### Full Blood Count

red blood cell count, white blood cell count, haemoglobin, haematocrit, mean cell volume, mean cell haemoglobin, mean cell haemoglobin concentration, red blood cell distribution width, platelet count, mean platelet volume, lymphocyte count, eosinophil count, neutrophil count, basophil count, monocyte count, lymphocyte %, eosinophil %, neutrophil %, basophil %, monocyte %

##### Liver Function Tests

alanine aminotransaminase, albumin, alkaline phosphatase, aspartate transaminase, bilirubin, alpha fetoprotein

##### Renal Function

sodium, potassium, creatinine, urea

##### Inflammatory Markers

C-reactive protein, erythrocyte sedimentation rate, plasma viscosity

### Other tests

amylase, HBA1c, calcium, calcium adjusted, total protein, blood glucose, fasting glucose, thyroid stimulating hormone

Data will also be extracted using SNOMED CT codes to explore the effect of the following covariates, which could independently affect the predictive value of blood test trends and likelihood of cancer:

1. Personal characteristics – age, sex, ethnicity, smoking history, alcohol intake, family/personal history of cancer, and patient-level Index of Multiple Deprivation (IMD) score.
2. Cancer symptoms and signs – symptoms shown to have an independent association with cancer as described by NICE^3^ and the primary care literature. All occurrences in the study period will be identified, allowing analyses of the first occurrence and repeat occurrences.
3. Results of other basic investigations available in primary care: B12 level, folate level, Carbohydrate Antigen 19-9 (CA19-9), Carcinoembryonic Antigen (CEA), Cancer Antigen 125 (CA125), Chest X-ray, Lactate Dehydrogenase (LDH), Prostate Specific Antigen (PSA), Iron Studies (Ferritin, Serum Iron, Total Iron Binding Capacity, Transferrin), Cholesterol.
4. Co-morbidity – recorded or implied from the prescribing record.
5. Prescribed medications.
6. Referrals for the urgent investigation of cancer, participation in cancer screening, and diagnosis route.

### Sample size

A sample size calculation was performed using the ‘*pmsampsize*’ package in Stata software, recently developed for prediction models by Riley *et al*^15^. The package uses the number of proposed predictor parameters in the model and the expected mean follow-up time, event rate, Cox-Snell R^2^, and amount of shrinkage (to adjust for overfitting). We based the calculation on our recent prediction model for 6-month risk of cancer following unexpected weight loss^12^. However, the appropriate risk window will be explored (Aim 3) so we performed the calculation twice: for 6-month risk and 1-year risk, to show the range of patient numbers required.

The number of proposed predictor variables in this study is 70, expected mean follow-up is 11 years (obtained from a second study using blood tests from CPRD^16^), 0.046 Cox-Snell R^2^ (from our recent model^12^) and expected 0.9 shrinkage factor. The 6-month event rate was 0.014 in our recent model (908 of 63,973 diagnosed^12^). We found no study that reported 1-year risk of any cancer in patients visiting primary care, so we used a conservative approach to obtain the 1-year event rate: as the outcome window is doubled from 6 months, we also doubled the event rate to give ∼1,800 events. However, patients are less likely to be diagnosed around the 1-year time-point so we decreased the number of events to 1,500. The 1-year event rate is then assumed 1500/63973 = 0.023. These inputs give 13,315 patients required at minimum. From these, we expect 146,465 person-years of follow-up, with 2,079 diagnosed in 6 months (∼30 events-per-variable) and 3,435 in 1 year (∼50 events-per-variable) (note: we request all patients, not 13,315 alone).

We expect to exceed these numbers, with another study reporting 69,942 incident cases of cancer within two years of reporting symptoms among 3,850,712 primary care patients^10, 11^. Another study using CPRD GOLD included all primary care patients available, reporting 10,875,556 full blood count blood tests among 1,893,641 primary care patients between 01/01/2000 and 14/01/2014^17^, providing reassurance that there will be sufficient data for analysis.

### Data/statistical analysis

Aim 1 (descriptive statistics):

1. Descriptive statistics will be used to describe patterns in blood testing, such as the frequency and time between serial tests, and summarise the results of each blood test overall and stratified by personal characteristics and comorbidity status. For the test results, descriptive statistics will include means (standard deviation) and medians (inter-quartile range). Associations between patient characteristics and patterns in blood testing will be examined.
2. The feasibility and comparison of established monitoring methods to summarise and identify blood test change over time (e.g. absolute change, percentage change, and rate of change).
3. Locally Weighted Scatterplot Smoothing (LOWESS) will be used to graphically describe trends in each blood test, summarised overall and by personal characteristics, cancer characteristics (diagnosis route, site, grade, histology, stage), and comorbidity status.
4. The mean (standard deviation) and median (inter-quartile range) number of days will be used to summarise the intervals between changes in blood test trends, cancer symptoms, single blood test abnormalities based on existing thresholds, referrals for cancer investigation or specialist review, and cancer diagnosis. Aim 2 (association with cancer):
5. Mixed-effects models: The trend in each blood level will be analysed using mixed-effects models. Multivariable models (one for each blood level) will include time (to model trends), personal, and clinical characteristics as fixed effects. Non-linearity will be assessed, such as non-linear relationships in blood levels over time. A random intercept for patient and random slope for time will be used to allow for differences in trends between patients. An unstructured correlation matrix will be used to account for repeated measures. Estimates will be presented with corresponding 95% confidence intervals (CIs). Lessons learned from this analysis, such as the relevant blood tests, confounders, and methods for handling non-linear trends, will inform the development of the prediction models in Aim 3.
6. Time-to-event model: Cox modelling will be used to establish temporal associations between personal and clinical characteristics and subsequent cancer diagnoses. Models will be repeated by cancer characteristics (diagnosis route, site, grade, histology, stage) to assess, for example, whether covariates are associated with individual or grouped cancer sites or can distinguish between early-stage and late-stage cancer. Estimates will be presented with corresponding 95% CIs. Lessons learned from this analysis will inform the development of the prediction models in Aim 3.
7. Joint models: Joint modelling of longitudinal blood test data and time-to-event data will be used for each blood level separately to assess the association between blood level trend and subsequent cancer diagnosis. In joint modelling, the trend is identified using a mixed-effects model and included as a covariate in the Cox model to assess association with subsequent cancer diagnosis. Initially, univariable joint models will be developed, which include only the trend (adjusted for personal and clinical characteristics, as per the mixed-effects model above). Multivariable joint models will then include the trend and personal and clinical characteristics related to cancer diagnosis (as per the Cox model above). Estimates will be presented with corresponding 95% CI. Lessons learnt from this analysis will inform the development of the prediction models in Aim 3.
8. Machine learning models, such as supervised clustering of trajectories using neural networks, will be considered to benchmark against the joint models. Instead of a traditional clustering method, which is unsupervised, the proposed method will cluster trends based on the outcome labels. As a result, clusters of different cancer characteristics (diagnosis route, site, grade, histology, stage) can be obtained, which allow us to provide patient-specific temporal association between outcome vs. exposure and covariates (such as symptoms and blood tests). Aim 3 (prediction):
9. Multivariate joint model: Multivariate joint modelling of longitudinal and time-to-event data will be developed to incorporate simultaneous trends in multiple blood tests to identify risk of cancer. The most promising blood test trends (as identified in the associations analysis described above) will be combined into a multivariate joint model, which will also include personal and clinical characteristics as covariates for subsequent cancer diagnosis. We will explore the predictive value of various clinical features and blood test trends, both as single covariates and in combinations. Pearson/Spearman correlation will be used to assess the correlation between blood components. Model coefficients will be presented with corresponding 95% CI.
10. Machine learning models, such as supervised clustering of trajectories using neural networks, will be considered to benchmark against multivariate joint modelling.
11. Model validation: Internal k-fold cross validation (where k = NHS geographical region) is planned to internally validate the model. However, this approach may not be possible due to the computational intensiveness of joint models. Alternatively, a split sample approach may be considered, whilst ensuring sample size requirements are met for model development. Performance statistics will be derived, such as sensitivity, specificity, predictive values, area under the receiver operating characteristic curve (or c-statistic), D-statistic for discrimination, Brier score, R^2^ statistic for explained variation, calibration slope, and calibration plots.
12. The chosen model(s) will be used to generate individual patient predictions. Predictions will be generated at multiple time-points along the blood test trajectory and performance measures derived for each time-point to identify when the optimal trend/threshold for referral for further investigations is reached.
13. Existing models incorporating blood test data to predict cancer diagnosis in primary care will be externally validated (by running the equations generated in the original derivation study on our CPRD data). Predictive performance will be compared to our new model(s). Performance of models incorporating blood test trend for cancer-risk will be compared to existing models including risk factors, symptoms, signs, static (or single) blood test data, and screening assessments. Performance measures described above will be used to assess diagnostic ability.
14. Decision curve analysis will be used to determine whether models incorporating blood test trend have superior clinical utility to models including static (or single) blood test data.

### Missing data

The amount of missing data and reasons, such as informative missingness and dropout, will be assessed for each blood test and other covariates. Levels of missing data will guide subsequent approaches to address missing data:

- Cox modelling - missing values for patient characteristics and blood test data will be replaced using a multiple imputation model including all predictors, the outcome, and auxiliary variables.
- Mixed-effects and joint modelling – due to the computationally intensive nature of mixed-effects and joint modelling and sporadic nature of blood testing over time, multiple imputation may not be possible. Therefore, we will initially model the data as-is and explore methods to account for missing longitudinal data.
- Machine learning – missing value imputation using a generative adversarial network will be explored and compared with the standard imputation methods in Cox.

### Patient and Public Involvement

We have set up a patient and public involvement (PPI) group, consisting of eight PPI advisors. The group will convene routinely throughout the project, sharing their experience with screening, symptoms, diagnostic pathways, treatment, outcome, and more. They will input into study dissemination.

## Discussion

In this paper, we describe the approved CPRD protocol designed to investigate blood test trends in patients attending primary care and the association between blood test trend(s) and diagnosis of cancer. The overall aim of BLOTTED is to explore whether incorporating blood test trends into the assessment of cancer risk in primary care may offer superior predictive performance to existing approaches to risk stratification. It is hypothesised that historical blood test trend may be used to trigger an urgent referral or direct access cancer investigation before symptoms develop or before individual blood test values reach an “actionable” threshold, as defined in current clinical guidance. We are particularly interested to understand whether the incorporation of blood test trajectory may increase the estimated risk of cancer sufficiently to trigger cancer investigation for patients with symptoms and risk factors that would not currently lead to a recommendation for cancer investigation. If patterns of blood test trend predict cancer in the absence of currently recognised risk factors and symptoms, an additional at-risk cohort may be identified for cancer investigation. The identification and referral of these additional patients may expedite their cancer diagnosis, reducing diagnostic delay. The individual and health system consequences of investigating these patients would merit further prospective and health economic evaluation.

### Limitations

A major limiting factor for BLOTTED is likely to be computational capacity. Employing advanced statistical modelling and machine learning to analyse longitudinal multivariable health records data from around 40 million patients in CPRD Aurum will lead to significant computational burden. We will work to develop and test our analytical approach on representative limited datasets before deploying the code on the main cohort. Additional technical resources and a secure centralised computing facility are also available to the research team at their host institution, if required to ensure the project is feasible.

A major consideration when developing an appropriate analytical approach will be to take into account the clinical indication for the available blood test data. Blood tests are ordered in routine clinical practice for many reasons besides cancer investigation. Patients without cancer may have another acute or chronic disease, or undergo treatment, that influence blood levels in a similar way to a malignant process. The frequency of testing is additionally dependent on a patient’s underlying consulting behaviour, their GPs clinical practice, and test access within the local health system.

Overall, the trends observed in the study population will represent a combination of testing driven by attendance, comorbidity, intra- and inter-person physiological variation over time, medication use, consultation and clinical behaviour. We plan to take into account for these factors during multivariable modelling so that false positives (patients determined to be high risk who are not at an increased risk of cancer) and false negatives (no discernible cancer signal despite high cancer risk) may be minimised.

### Implications for clinical practice

There are currently no diagnostic prediction models that utilise available data by incorporating blood test trends integrated into the electronic health record. Therefore, a vast amount of historical information is missing from approaches to risk stratification in primary care. Alongside the identification of promising discriminative approaches to risk stratification that incorporate blood test trend, we will conduct future research to understand the implementation challenges of integrating them in clinical practice and explore whether there are existing integrated approaches that could be modified to incorporate our findings. If methods require significant processing power, significant technological development will be required to enable timely actionable risk stratification in the clinic. Once available in clinical practice, new research will be required to understand the implementation challenges, not least GP uptake and patient acceptance of this method of patient selection.

## Data Availability

Not applicable, as this is a study protocol

## Abbreviations

(CA125): Cancer Antigen 125
(CA19-9): Carbohydrate Antigen 19-9
(CEA): Carcinoembryonic Antigen
(CPRD): Clinical Practice Research Datalink
(CI): Confidence interval
(GP): General practitionor
(HES): Hospital Episode Statistics
(IMD): Index of Multiple Deprivation
(LDH): Lactate Dehydrogenase
(LOWESS): Locally Weighted Scatterplot Smoothing
(MDC): Multidisciplinary Diagnostic Centre
(NCRAS): National Cancer Registration and Analysis Service
(NHS): National Health Service
(NICE): National Institute for Health and Care Excellence
(ONS): Office of National Statistics
(PPI): Patient and public involvement
(PSA): Prostate Specific Antigen

## Additional information

### Ethical approval and consent

CPRD has ethical approval from the Health Research Authority to hold anonymised patient data and to support research using that data. CPRD’s approval of data access for individual research projects includes ethics approval and consent for those projects. Ethical approval will therefore be covered for this study by the CPRD.

### Consent for publication

Not applicable

### Availabbility of data and materials

Not applicable

## Author contributions

PSV and BDN wrote the first draft of the protocol. All authors reviewed and updated draft versions and contributed to revisions. All authors approved the final manuscript.

## Acknowledgements

The authors would like to thank PPI representatives Bernard Gudgin, Julian Ashton, Clara Martins de Barros, Shannon Draisey, Susan Lynne, Emily Lam, Ian Blelloch, and Margaret Ogden for input into protocol development.

## References

[1] Watson J, Mounce L, Bailey SE, Cooper SL, Hamilton W. Blood markers for cancer. BMJ (Clinical researched). 2019;367:5774.

[2] Rubin GP, Saunders CL, Abel GA, McPhail S, Lyratzopoulos G, Neal RD. Impact of investigations in general practice on timeliness of referral for patients subsequently diagnosed with cancer: analysis of national primary care audit data. Br J Cancer. 2015;112(4):676–87.

[3] NICE. Suspected cancer: recognition and referral (NG12). National Institute for Health and Care Excellence 2015. Available online at <https://www.nice.org.uk/guidance/ng12>. Last Accessed 4th March 2022.

[4] Bull LM, Lunt M, Martin GP, Hyrich K, Sergeant JC, Harnessing repeated measurements of predictor variables for clinical risk prediction: a review of existing methods. Diagn Progn Res, 2020. 4: p. 9.

[5] Virdee PS., Patnick P, Watkinson P, Birks J, Holt T. Trends in the full blood count blood test and colorectal cancer detection: a longitudinal, case-control study of UK primary care patient data. NIHR Open Research, 2022, 2, 32:1–53. DOI: 10.3310/nihropenres.13266.1.

[6] Virdee PS, Birks J, Holt T. A dynamic prediction model for early detection of colorectal cancer using routine blood test results from primary care. SAPC ASM 2021 - virtual conference. Available online at <https://sapc.ac.uk/conference/2021/abstract/dynamic-prediction-model-early-detection-of-colorectal-cancer-using-routine>. Last Accessed 4th March 2022.

[7] Nicholson BD, Perera R, Thompson MJ. The elusive diagnosis of cancer: testing times. Br J Gen Pract. 2018;68(676):510–1.

[8] Koshiaris C, Van den Bruel A, Oke JL, Nicholson BD, Shephard E, Braddick M, et al. Early detection of multiple myeloma in primary care using blood tests: a case-control study in primary care. Br J Gen Pract. 2018;68(674):e586–e93.

[9] Naeser E, Moller H, Fredberg U, Frystyk J, Vedsted P. Routine blood tests and probability of cancer in patients referred with non-specific serious symptoms: a cohort study. BMC Cancer. 2017;17(1):817.

[10] Hippisley-Cox J, Coupland C. Symptoms and risk factors to identify women with suspected cancer in primary care: derivation and validation of an algorithm. Br J Gen Pract. 2013;63(606):e11–21.

[11] Hippisley-Cox J, Coupland C. Symptoms and risk factors to identify men with suspected cancer in primary care: derivation and validation of an algorithm. Br J Gen Pract. 2013 Jan;63(606):e1–10.

[12] Nicholson BD, Aveyard P, Koshiaris C, Perera R, Hamilton W, et al. Combining simple blood tests to identify primary care patients with unexpected weight loss for cancer investigation: Clinical risk score development, internal validation, and net benefit analysis. PLOS Medicine. 2021;18(8):e1003728.

[13] Clinical Practice Research Datalink (CPRD). 2022 [Accessed 25 August 2022]; Available from https://www.cprd.com/.

[14] Virdee PS, Fuller A, Jacobs M, Holt T, Birks J, Assessing data quality from the Clinical Practice Research Datalink: a methodological approach applied to the full blood count blood test. J big Data, 2020. 7, 95:1–18. DOI: 10.1186/s40537-020-00375-w.

[15] Riley, R.D., Ensor, J., Snell, K.I.E., Harrell, F.E., Jr., Martin, G.P., Reitsma, J.B., Moons, K.G.M., Collins, G., and van Smeden, M., Calculating the sample size required for developing a clinical prediction model. BMJ. 2020;368:p. m441

[16] Birks J, Bankhead C, Holt T, Fuller A, Patnick J. Evaluation of a prediction model for colorectal cancer: retrospective analysis of 2.5 million patient records. Cancer Med. 2017 Oct;6(10):2453–2460.

[17] Holt T, Birks J, Bankhead C, Nicholson BD, Fuller A, Patnick J. Do changes in full blood count indices predate symptom reporting in people with undiagnosed bowel cancer? Retrospective analysis using cohort and case control designs. SAPC ASM 2021 - virtual conference. Available online at <https://sapc.ac.uk/conference/2021/abstract/do-changes-full-blood-count-indices-predate-symptom-reporting-people>. Last Accessed 4th March 2022.

